# Clinical prediction models for the management of blunt chest trauma in the Emergency Department: a systematic review

**DOI:** 10.1101/2023.06.15.23291429

**Authors:** Ceri Battle, Elaine Cole, Kym Carter, Edward Baker

## Abstract

**Introduction:** The aim of this systematic review was to investigate how clinical prediction models compare in terms of their methodological development, validation, and predictive capabilities, for patients with blunt chest trauma presenting to the Emergency Department.

**Methods:** A systematic review was conducted across databases from Jan 2000 until March 2023. Studies were categorised into three types of multivariable prediction research and data extracted regarding methodological issues and the predictive capabilities of each model. Risk of bias and applicability were assessed.

**Results:** 39 studies were included that discussed 22 different models. The most commonly observed study design was a single-centre, retrospective, chart review. The most widely externally validated clinical prediction models with moderate to good discrimination were the Thoracic Trauma Severity Score and the STUMBL Score.

**Discussion:** This review demonstrates that the predictive ability of some of the existing clinical prediction models is acceptable, but high risk of bias and lack of subsequent external validation limits the extensive application of the models. The Thoracic Trauma Severity Score and STUMBL Score demonstrate better predictive accuracy in both development and external validation studies than the other models, but require recalibration and / or update and evaluation of their clinical and cost effectiveness.

## INTRODUCTION

Patients with blunt chest trauma present an ongoing challenge for accurate triage in the Emergency Department (ED). Whilst the majority of patients with blunt chest trauma will have an uncomplicated recovery, clinical presentation at the time of ED assessment is no guarantee that a patient will be of suitable acuity for discharge to home, or for admission to award setting, as up to 10% of patients will decompensate after 48-72 hours.^1–3^ Intensive Care Unit (ICU) referral from the ED must be carefully considered and as a result, much has been published over the last 20 years investigating the predictors of poor outcome in this patient cohort.^4, 5^ These predictors include patient age, severity of injury, number and location of rib fractures, pre-injury anticoagulants, chronic lung disease and others.^4, 6–8^

A common aim of such primary prognostic studies is the development of clinical prediction models. The clinical prediction model is intended to estimate the individualised probability or risk that a condition, for example mortality or pulmonary complications, will occur in the future by combining multiple prognostic factors / predictors from an individual.^9, 10^ A number of different clinical prediction models have been developed for patients with blunt chest trauma, however there is still no universally accepted model in clinical practice. A recent survey study highlighted that there were 20 different clinical prediction models and pathways used when assessing whether a patient with blunt chest trauma is safe for ED home discharge.^11^

There is often conflicting evidence regarding the predictive capabilities of developed clinical prediction models, leading to a growing demand for evidence synthesis of external validation studies that assess model performance in a new patient cohort.^10, 12, 13^ This is applicable to the range of clinical prediction models used for the management of patients with blunt chest trauma. The aim of this systematic review therefore was to investigate how clinical prediction models compare in terms of their methodological development, validation, and predictive capabilities, for clinical and healthcare utilisation outcomes for patients with blunt chest trauma presenting to the Emergency Department.

## METHODS

### Search strategy

The review was registered prospectively on the PROSPERO database (https://www.crd.york.ac.uk/PROSPERO/display_record.php?RecordID=351638). The CHARMS Checklist was followed for completion of this review. A broad search strategy was employed in order to capture all relevant studies. The search filter was used for PubMed and Embase Databases, the Cochrane Library, and OpenGrey from Jan 2000 until March 2023.

The search term combinations were based on Geersing et al (2012)^12^ and used Medical Subject Heading terms, text words and word variants for blunt chest trauma. These were combined with relevant terms for both outcomes and clinical prediction model development and validation methods. The search strategy can be found in online supplementary file 1. The reference lists of all relevant studies were hand-searched in order to identify any evidence missed in the electronic search. The Annals of Emergency Medicine, Emergency Medicine Journal, Injury and the Journal of Trauma and Acute Care Surgery were hand-searched for relevant studies. Searches were international and no search limitations were used.

### Study selection

Studies were included that focussed on patients aged ≥16 presenting to the Emergency Department with blunt chest trauma (defined as a blunt chest injury resulting in chest wall contusion or rib fractures, with or without underlying lung injury). Prognostic multivariable prediction studies were included where the aim of the study was to predict an outcome using two or more independent variables, in order to develop a multivariable (at least two variables) weighted clinical prediction model for any outcome following blunt chest trauma. Based on the ‘Critical Appraisal and Data Extraction for Systematic Reviews of Prediction Modelling Studies: CHARMS guidance^13^, studies were categorised into three types of multivariable prediction research; 1) model development studies without external validation. 2) model development studies with external validation in independent data, and 3) external validation studies without or with model updating.

Studies were excluded which included patients presenting with: a) Penetrating trauma only, b) Multi-trauma only and no reference to chest trauma, c) Severe intra-thoracic injuries only (eg. bronchial, cardiac, oesophageal, aortic or diaphragmatic rupture) and no chest wall trauma, d) Children aged <16 years. Other exclusion criteria included, studies that investigated a single predictor (such as single prognostic marker studies), studies that investigated only causality between one or more variables and an outcome, and studies that do not contribute to patient care. For multiple publications from the same dataset, only the most relevant study to this reviews aims was included. Studies for which only an abstract was available were also excluded.

### Data extraction

A two-step process was used to reduce potential selection bias. Two researchers (CB and EB) analysed each title and abstract independently and then met to discuss any discrepancies. The full paper of selected studies was analysed by the reviewers. Data were extracted relating to both the reporting of and use of methods known to influence the quality of multivariable prediction studies. A data extraction form based on CHARMS Checklist was used to record relevant information, a copy of which is available in supplementary file 2[. Study authors were contacted for any missing data and response time set at six weeks. Included studies were grouped according to the clinical prediction model under investigation for the analysis.

Data were extracted regarding the methodological issues that are considered to be important in prediction research, focussed broadly on the reporting of the domains outlined in the CHARMS Checklist. Data regarding the predictive capabilities of each model were also extracted where available, for the following outcomes; a) clinical outcomes such as mortality and any pulmonary complications, and b) healthcare utilisation outcomes such as length of stay, need for mechanical ventilation or ICU admission.

### Quality assessment

Risk of bias and applicability were assessed using the “Prediction model Risk Of Bias ASsessment Tool” (PROBAST)^14^ where: “Risk of bias refers to the extent that flaws in the design, conduct, and analysis of the primary prediction modelling study lead to biased, often overly optimistic, estimates of predictive performance measures such as model calibration, discrimination, or (re)classification (usually due to over-fitted models). Applicability refers to the extent to which the primary study matches the review question, and thus is applicable for the intended use of the reviewed prediction model(s) in the target population” (Moons et al, 2014). PROBAST includes 20 signalling questions across four domains (participants, predictors, outcome, and analysis) which were scored low, high or unclear. For each included study, an overall final score for judgement of risk of bias and applicability was allocated. This process was completed independently by two reviewers (CB and EB), with a third reviewer (EC) used to resolve any discrepancies. An outline of the PROBAST Score can be found in online supplement 3.

### Data synthesis and analysis

Narrative synthesis of included study results was conducted, grouped according to clinical prediction models. Model performance was evaluated through assessment of model discrimination, a measure of how well the model can separate those who do and those who do not have the disease of interest, and calibration, a measure of how well predicted probabilities agree with the actual observed risk. The discrimination ‘C-statistic’ (balance between negative and positive predictive value) was defined as low (below 0⋅70), moderate (0⋅70–0⋅79) or good (at least 0⋅80). Where available in the studies, the correlation between observed and expected (calibration) outcome, as measured by the Hosmer–Lemeshow (H-L) test, was presented using a p >0⋅050 to indicate a good model fit.^13^

## RESULTS

### Study selection

The initial search strategy identified 9495 citations. Following screening titles and abstracts, we identified 172 potentially relevant studies and following full-text review, a total of 39 studies met the inclusion criteria. No additional citations were identified through the grey literature or reference list searches. Figure 1 outlines the flow diagram of study selection.

**Figure 1:**
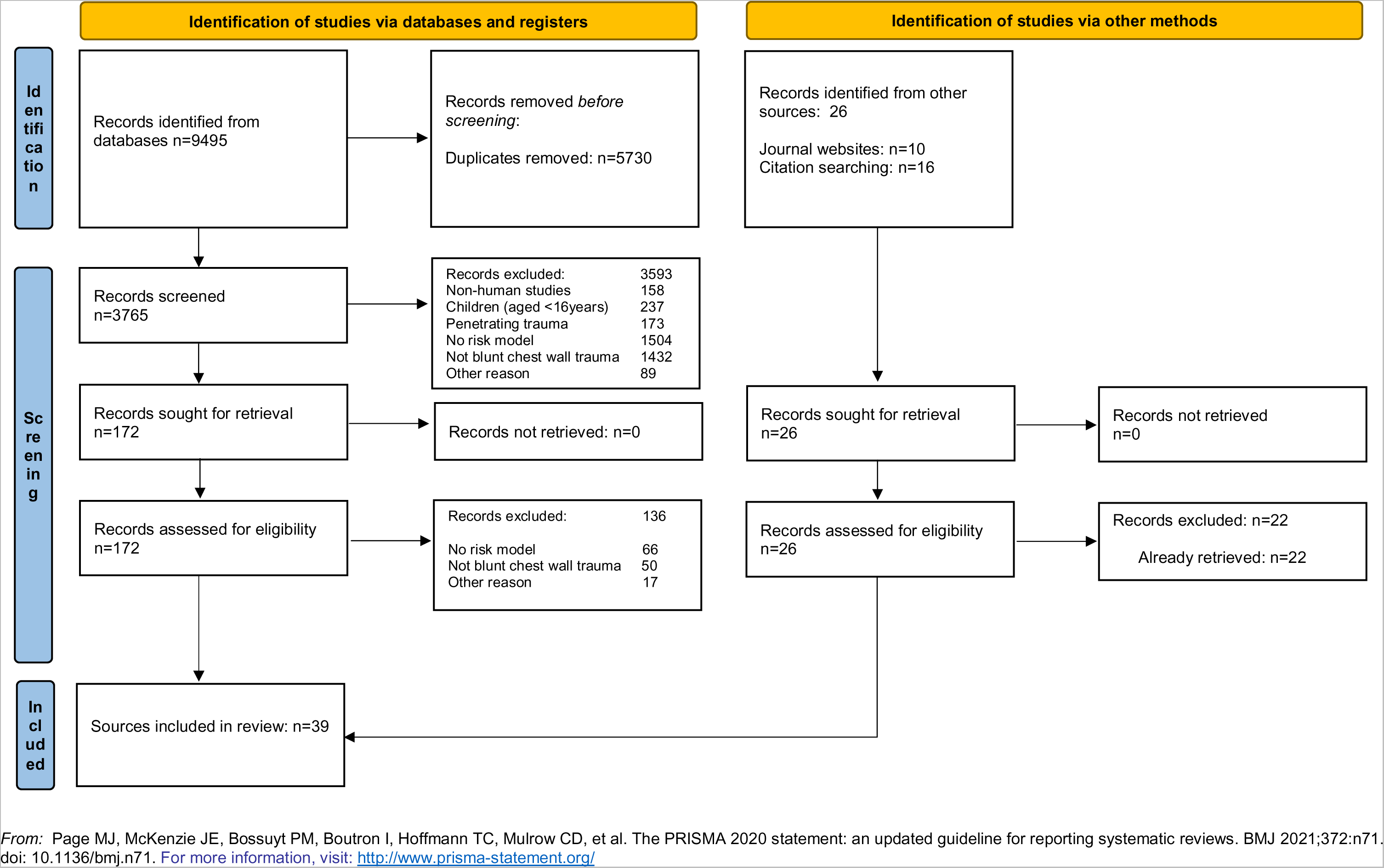
PRISMA Flow Diagram

### Study characteristics

The 39 studies were categorised as; 12 model development studies without external validation, three model development studies with external validation in independent data, and 24 external validation studies without or with model updating. The most commonly observed study design was a single-centre, retrospective, chart review. A total of 22 different clinical prediction models were studied and therefore included in this review. Study design, clinical prediction model, study population (including diversity data where possible, such as age, sex, frailty and ethnicity), total sample size, outcomes and results of the included studies are outlined in Table 1.

**Table 1:**
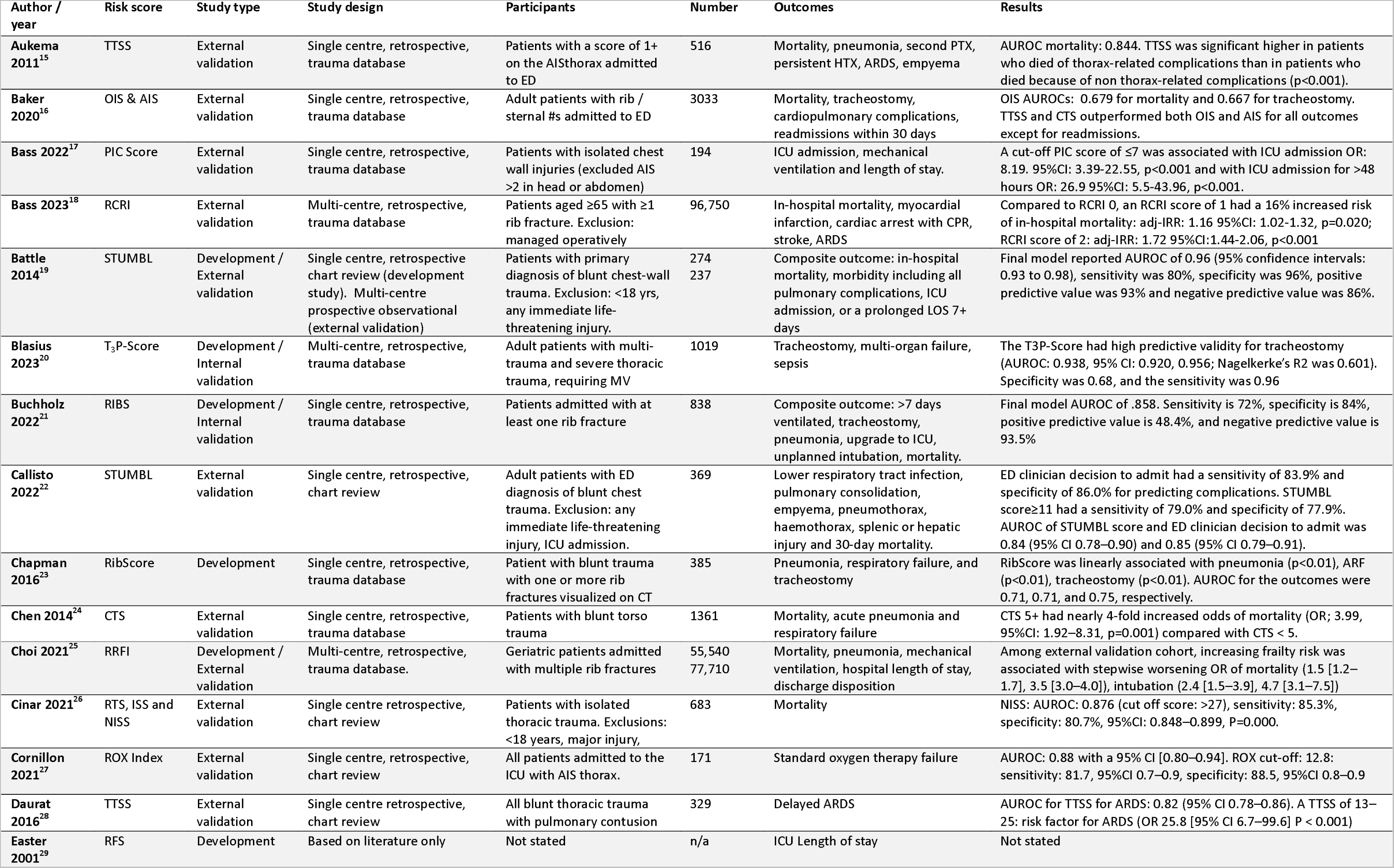
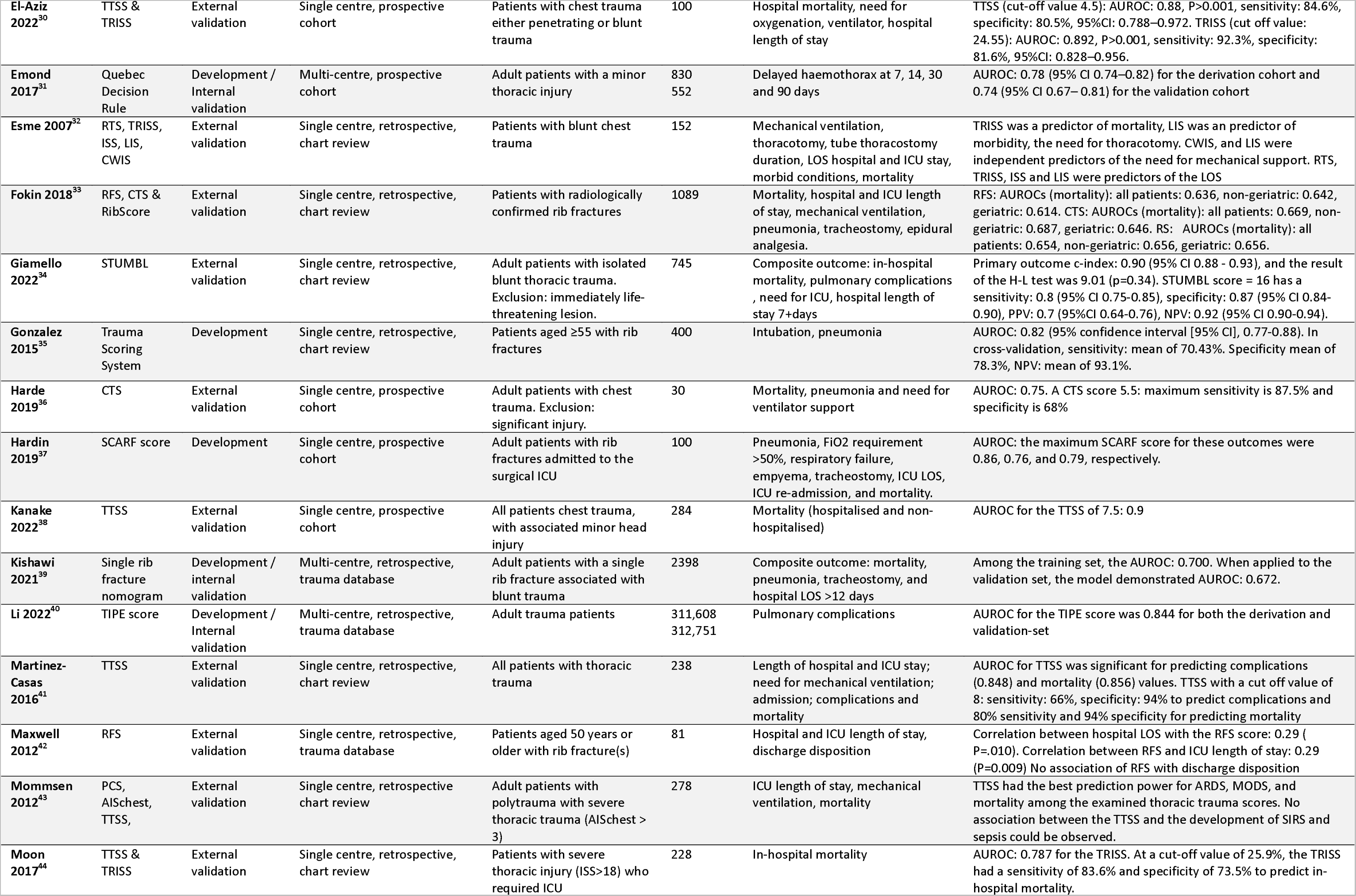
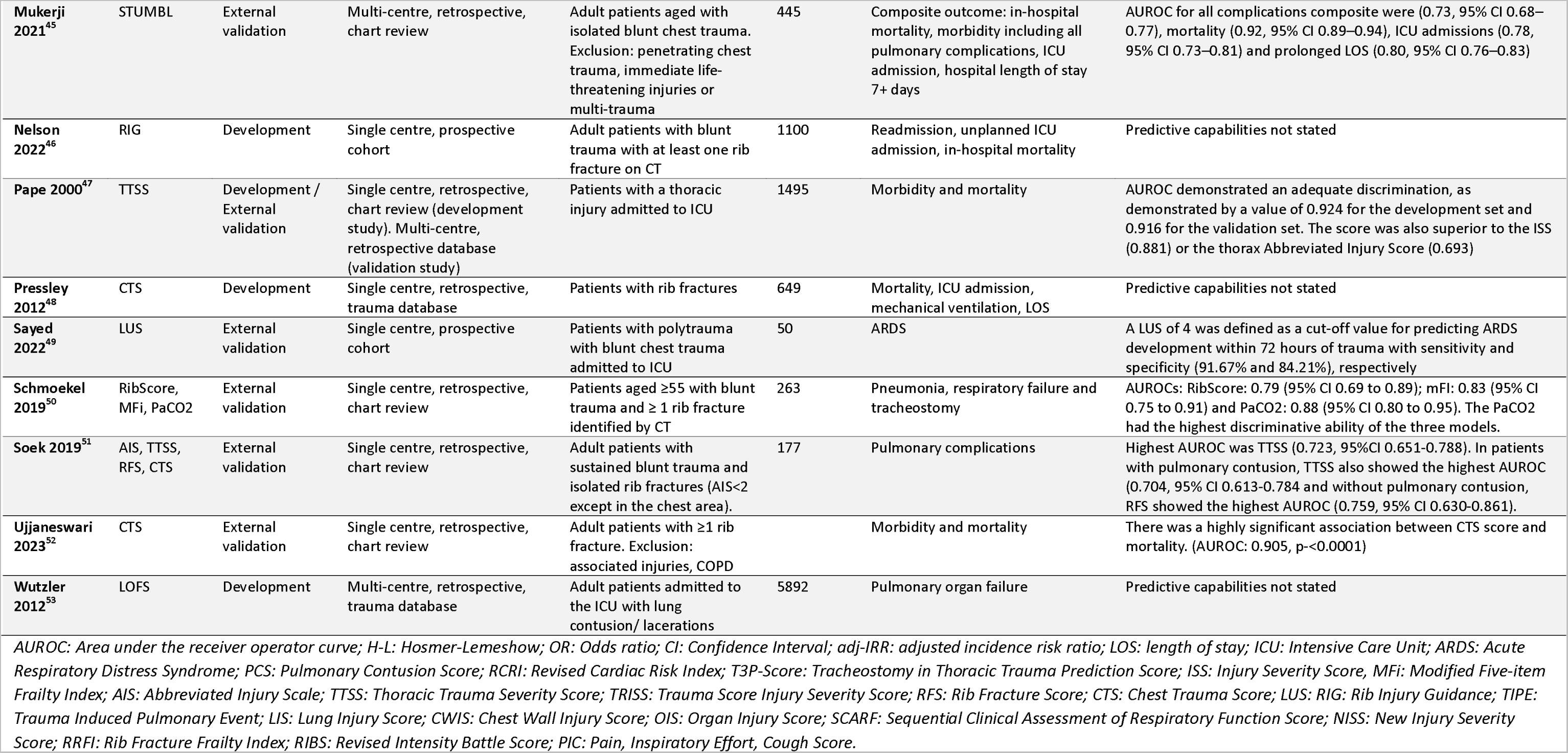
Characteristics of included studies

### Quality assessment

The quality of the included studies in this review was variable. Risk of bias was high across most of the included studies for the analysis. Selection of predictors was commonly based on univariable analysis result, handling of missing data was inadequately described and the model performance measures, in particular the model’s calibration, was infrequently reported. The studies scored mostly low risk of bias in terms of the predictors included. Risk of bias for participants was variable across the studies as some used a trauma registry for their participant data. In terms of applicability, some studies scored high risk for participants, as they included paediatric patients, which this review was not investigating. The full PROBAST results are outlined in Table 2 and Figure 2.

**Figure 2:**
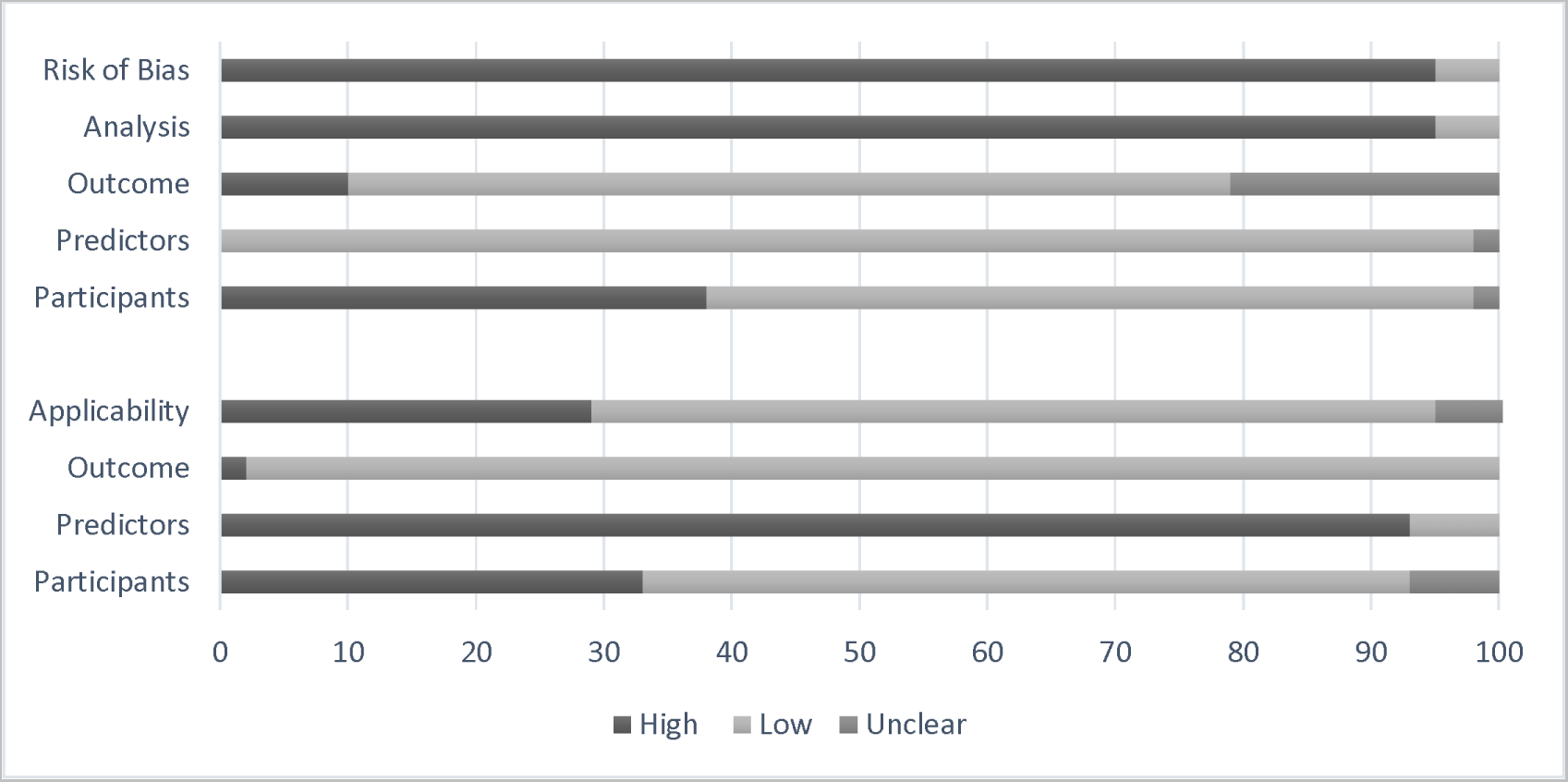
Risk of bias and applicability of included studies: PROBAST results

**Table 2:**
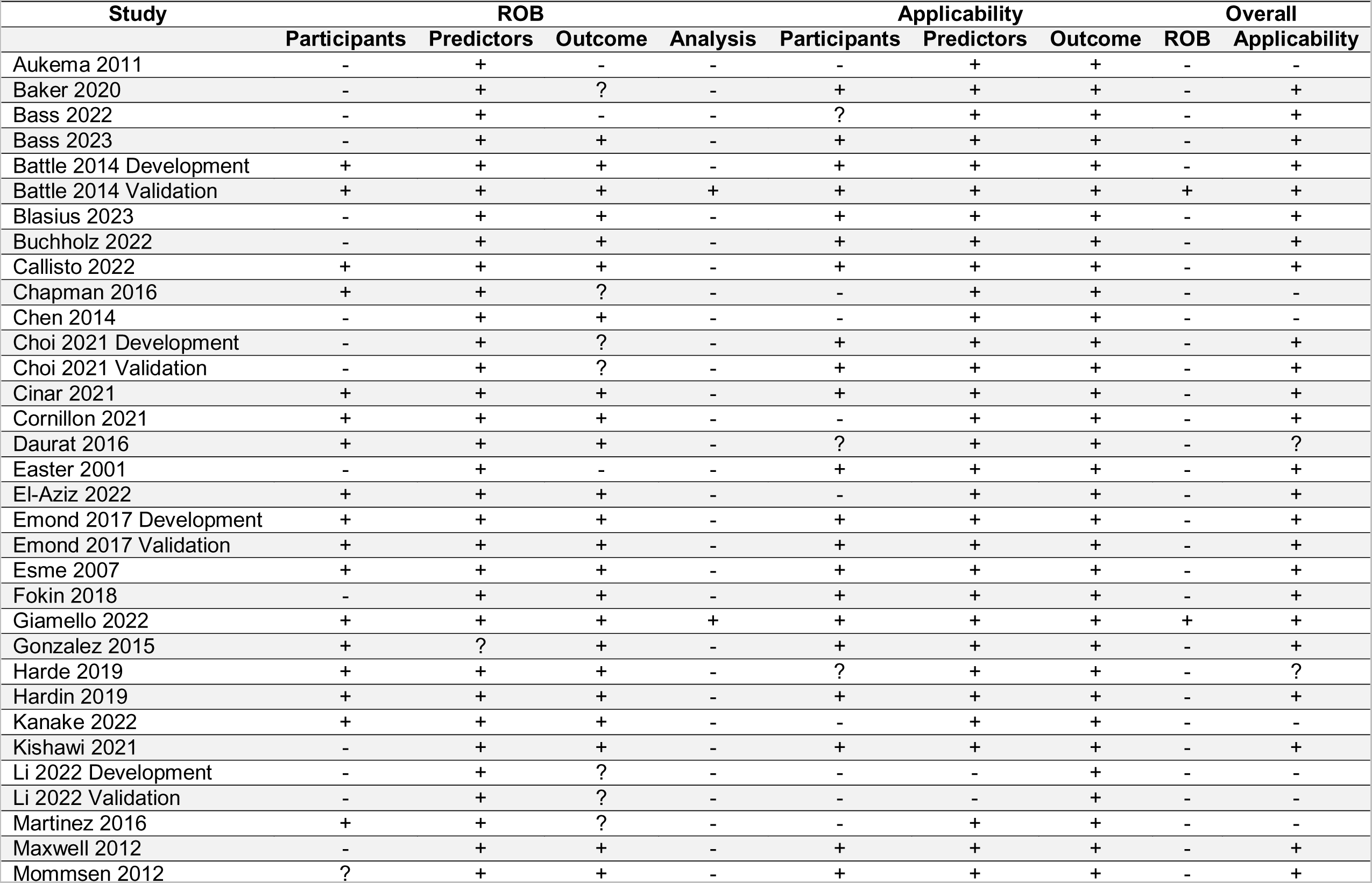
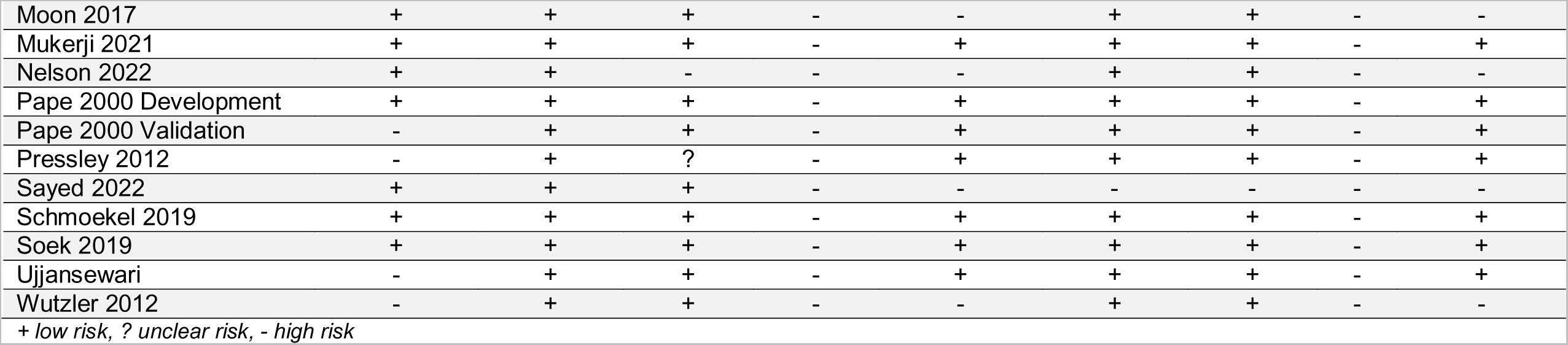
Risk of bias and applicability of included studies: PROBAST results

| Study | ROB |  |  |  | Applicability |  |  | Overall |  |
| --- | --- | --- | --- | --- | --- | --- | --- | --- | --- |
|  | Participants | Predictors | Outcome | Analysis | Participants | Predictors | Outcome | ROB | Applicability |
| Aukema 2011 | - | + | - | - | - | + | + | - | - |
| Baker 2020 | - | + | ? | - | + | + | + | - | + |
| Bass 2022 | - | + | - | - | ? | + | + | - | + |
| Bass 2023 | - | + | + | - | + | + | + | - | + |
| Battle 2014 Development | + | + | + | - | + | + | + | - | + |
| Battle 2014 Validation | + | + | + | + | + | + | + | + | + |
| Blasius 2023 | - | + | + | - | + | + | + | - | + |
| Buchholz 2022 | - | + | + | - | + | + | + | - | + |
| Callisto 2022 | + | + | + | - | + | + | + | - | + |
| Chapman 2016 | + | + | ? | - | - | + | + | - | - |
| Chen 2014 | - | + | + | - | - | + | + | - | - |
| Choi 2021 Development | - | + | ? | - | + | + | + | - | + |
| Choi 2021 Validation | - | + | ? | - | + | + | + | - | + |
| Cinar 2021 | + | + | + | - | + | + | + | - | + |
| Cornillon 2021 | + | + | + | - | - | + | + | - | + |
| Daurat 2016 | + | + | + | - | ? | + | + | - | ? |
| Easter 2001 | - | + | - | - | + | + | + | - | + |
| El-Aziz 2022 | + | + | + | - | - | + | + | - | + |
| Emond 2017 Development | + | + | + | - | + | + | + | - | + |
| Emond 2017 Validation | + | + | + | - | + | + | + | - | + |
| Esme 2007 | + | + | + | - | + | + | + | - | + |
| Fokin 2018 | - | + | + | - | + | + | + | - | + |
| Giamello 2022 | + | + | + | + | + | + | + | + | + |
| Gonzalez 2015 | + | ? | + | - | + | + | + | - | + |
| Harde 2019 | + | + | + | - | ? | + | + | - | ? |
| Hardin 2019 | + | + | + | - | + | + | + | - | + |
| Kanake 2022 | + | + | + | - | - | + | + | - | - |
| Kishawi 2021 | - | + | + | - | + | + | + | - | + |
| Li 2022 Development | - | + | ? | - | - | - | + | - | - |
| Li 2022 Validation | - | + | ? | - | - | - | + | - | - |
| Martinez 2016 | + | + | ? | - | - | + | + | - | - |
| Maxwell 2012 | - | + | + | - | + | + | + | - | + |
| Mommsen 2012 | ? | + | + | - | + | + | + | - | + |
| Moon 2017 | + | + | + | - | - | + | + | - | - |
| Mukerji 2021 | + | + | + | - | + | + | + | - | + |
| Nelson 2022 | + | + | - | - | - | + | + | - | - |
| Pape 2000 Development | + | + | + | - | + | + | + | - | + |
| Pape 2000 Validation | - | + | + | - | + | + | + | - | + |
| Pressley 2012 | - | + | ? | - | + | + | + | - | + |
| Sayed 2022 | + | + | + | - | - | - | - | - | - |
| Schmoekel 2019 | + | + | + | - | + | + | + | - | + |
| Soek 2019 | + | + | + | - | + | + | + | - | + |
| Ujjansewari | - | + | + | - | + | + | + | - | + |
| Wutzler 2012 | - | + | + | - | - | + | + | - | - |

+ low risk, ? unclear risk, - high risk

Figure 2 demonstrates the overall judgment of the included studies.

### Clinical prediction models

#### Thoracic Trauma Severity Score (TTSS)

The TTSS was originally developed and externally validated by Pape et al (2000) to predict the risk of thoracic-trauma related complications in patients with blunt polytrauma, admitted to ICU.^47^ Based on high risk of bias results, the c-index demonstrated good discrimination, as demonstrated by a value of 0.924 for the development set and 0.916 for the validation set, although 95% confidence intervals were not reported. Since 2002, there have been nine external validation studies of high risk of bias, that have reported various cut off values on the TTSS, with moderate to good level c-indices ranging between 0.723 and 0.848. Model calibration was not reported in any of the included studies.

#### STUMBL Score

The STUMBL Score was original developed and externally validated by Battle et al (2014) to predict risk of pulmonary complications in patients with isolated blunt chest wall trauma presenting to the ED.^19^ Based on low risk of bias results, the final model demonstrated good discrimination with a reported c-index of 0.96 (95% CI: 0.93 to 0.98). The model showed good calibration when evaluated with the Hosmer Lemeshow test (9.22, P = 0.32). Since development, there have been three external validation studies completed of variable risk of bias, that have reported various cut off values on the STUMBL Score, with moderate to good level c-indices ranging between 0.73 (95% CI 0.68–0.77) and 0.90 (95% CI 0.88 - 0.93).

#### Rib Fracture Score (RFS)

The RFS was originally developed by Easter et al (2001), as a protocol for the management of pain, respiratory care and mobility in patients with multiple rib fractures.^29^ The score allocated to the patient (based on number of fractures, number of sides and the patient’s age), determines the treatment recommendations, rather than a risk of a particular outcome. The protocol was based on literature, rather than patient data and as a result was at high risk of bias. No predictive capabilities were reported in the original development study. Three external validation studies of high risk of bias, have been completed, demonstrating a low level of discrimination with c-indices ranging from 0.64 to 0.67 for the prediction of a number of clinical and healthcare resource outcomes. Model calibration was not reported in the included studies.

#### Chest Trauma Score (CTS)

The CTS, originally developed by Pressley et al (2012) for patients presenting with rib fractures, using clinical data available at the time of initial evaluation. It predicts the likelihood of mechanical ventilation and prolonged courses of care.^48^ The development study does not report predictive capabilities of the score and was considered high risk of bias. Five external validation studies of high risk of bias have been completed, demonstrating a low to good level of discrimination with c-indices of 0.67 to 0.91. Model calibration was not reported in any of the studies.

#### RibScore

The RibScore, originally developed by Chapman et al (2016) for blunt trauma patients with rib fractures, was based on six candidate radiographic variables, identified on CT imaging.^23^ They reported c-indices the outcomes pneumonia, respiratory failure and tracheostomy were 0.71, 0.71, and 0.75, respectively in a high risk of bias study. Two high risk of bias external validation studies have been completed in which low and moderate c-indices of 0.66 and 0.79 (95% CI 0.69 to 0.89) were reported. Model calibration was not reported in any of the studies.

#### Other clinical prediction models

Table 1 outlines 18 other clinical prediction models which were identified, for which only one study (all high risk of bias) per model met the inclusion criteria for this review. A number of new clinical prediction models have been developed (all high risk of bias studies) but not yet validated were included in the review. These included the Tracheostomy in Thoracic Trauma Prediction Score^20^ (T_3_P-Score, c-index for tracheostomy: 0.938, 95% CI: 0.920-0.956), Sequential Clinical Assessment of Respiratory Function^37^ (SCARF Score, c-index for pneumonia: 0.86), Rib Injury Guidelines^46^ (RIG, c-index not reported), the Lung Organ Failure Score^53^ (c-index not reported), and a new scoring system^35^ (c-index: 0.82; 95% CI: 0.77-0.88).

Other models developed and validated by the original authors, but yet to be externally validated in further studies included The Rib Fracture Frailty Index^25^ (RFFI) (c-index not reported), the Revised Intensity Battle Score^21^ (RIBS) (c-index: 0.86), Quebec Minor Thoracic Injury Decision Rule^31^ (c-index: 0.78; 95% CI 0.74–0.82), a single rib fracture nomogram^39^ (c-index: 0.70), and the Trauma Induced Pulmonary Event (TIPE Score) (c-index: 0.85).^40^

The chest wall components of the Abbreviated Injury Scale (AIS) and Organ Injury Scale (OIS) were externally validated in a high risk of bias study by Baker et al (2020) which reported a low level of discrimination for both the OIS (c-index: 0.68; 95% CI: 0.64-0.73) and AIS (c-index: 0.59; 95%CI: 0.55 to 0.63) for patients with rib and sternal fractures presenting to the ED.^16^

There were four model development studies that did not meet the inclusion criteria for this review, but subsequent validation studies were included (all high risk of bias). These included the Revised Cardiac Risk Index^18^ (RCRI, originally developed to predict 30-day postoperative myocardial infarction, cardiac arrest, or mortality following non-cardiac surgery, c-index not reported), Pain Inspiratory Effort Cough Score^17^ (PIC Score, c-index not reported), Revised Trauma Scale^26^ (RTS, c-index: 0.76, 95%CI: 0.72-0.79), Lung Ultrasound Score^49^ (LUS, c-index not reported), and the ROX Index^27^ (which combines respiratory rate and oxygenation values, c-index: 0.88; 95%CI: 0.80–0.94).

## DISCUSSION

This systematic review has highlighted that there are numerous clinical prediction models used for the management of patients with blunt chest trauma in various healthcare settings. These models differ widely in terms of their target patient population, included risk factors and outcomes predicted. They also differ in terms of the methods used for both their development and validation. These findings impede comparison between the models and generalisability for the patient with blunt chest wall trauma. These inherent differences also contribute to the lack of consensus in clinical practice, regarding the optimal clinical prediction model for this patient population.^54, 55^

This review highlights the difficulties in developing, validating and using a clinical prediction model. Instead of updating existing models and improving their predictive capabilities, most studies have developed and presented a new model. This has resulted in better performance in their population compared with existing models that were developed in another population and validated externally. Furthermore, there were no impact studies retrieved in this review that explored the clinical or cost effectiveness of any of the models. Traditional impact studies are reported to be costly to undertake and as a result, very few exist for any patient condition.^55^ It is reasonable therefore to suggest that the ideal model does not yet exist.

Not all studies calculated a c-index to describe the discriminative abilities of the model and only one study reported a H-L analysis for calibration. Other studies may have used alternative measurements, or it must be assumed that they have compared observed with expected results, but did not report the comparison statistic. Overall, discrimination is more straightforward to calculate when compared with calibration, and the latter can be easily improved using updating methods applied to a new patient cohort.^13, 55^ Good calibration is necessary however for calculating predictions, independent of the reported c-index.^55^ The clinical usefulness of a model can only be determined when both discrimination and calibration are available, and a model’s cut-off value has been defined for reported sensitivity and specificity values.^13, 55^

The most widely externally validated clinical prediction models with moderate to good discrimination developed specifically for the management of patients with blunt chest trauma, were the TTSS^47^ and STUMBL Score^19^. These models were developed for use in different healthcare settings and only the STUMBL Score had been assessed for calibration. Neither model has undergone any recalibration, updating or revision, nor have been assessed for clinical or cost effectiveness. There is limited reference to different diverse patient groups in any of the included studies, with exception to the STUMBL Score, which was the only model that was reported to have been specifically externally validated on patients of varying ethnic groups. Health inequalities across ethnic groups are reported in other disease populations^56, 57^ but currently it isn’t clear if existing blunt chest trauma clinical prediction models account for diversity-related differences.

This systematic review has a number of limitations. A large number of the included studies failed to report confidence intervals for the reported c-indices, resulting in incomplete comparisons between the models. Most of these models had been developed on Causcian populations, and it remains unknown (other than the STUMBL Score New Zealand validation study^45^) whether these models would perform equally well in other ethnic groups. Frailty as a potential candidate predictor was not considered in any of the included model development studies, other than the RFFI study.^25^ It is well-recognised that frailty identification has an important role in any clinical decision-making related in older trauma patients^58, 59^, therefore this needs further consideration in future studies and existing model updates. Finally, the lead author of this review is also the researcher who developed the STUMBL Score, so there is the potential for interpretive bias.

In conclusion, this systematic review has examined the methodological development, validation, and predictive capabilities of the clinical prediction models, for clinical and healthcare utilisation outcomes for patients with blunt chest trauma presenting to the Emergency Department. The predictive ability of some of the existing clinical prediction models is acceptable, but high risk of bias and lack of subsequent external validation limits the extensive application of the models in the general blunt chest trauma population. TTSS and STUMBL Score demonstrate better predictive accuracy in both development and external validation studies than the other models, but both potentially still require recalibration and / or update and evaluation of their clinical and cost effectiveness.

## Data Availability

All data produced in the present study are available upon reasonable request to the authors

