## Supplementary file 1 for "Clinical prediction models for the management of blunt chest trauma in the Emergency Department: a systematic review"

**Search strategy example PUBMED**

chest trauma[title/abstract] AND stratification[title/abstract]

chest trauma[title/abstract] AND ROC curve[title/abstract]

chest trauma[title/abstract] AND discriminat*[title/abstract]

chest trauma[title/abstract] AND c statistic[title/abstract]

chest trauma[title/abstract] AND area under the curve[title/abstract]

chest trauma[title/abstract] AND AUC[title/abstract]

chest trauma[title/abstract] AND calibration[title/abstract]

chest trauma[title/abstract] AND indices[title/abstract]

chest trauma[title/abstract] AND algorithm[title/abstract]

chest trauma[title/abstract] AND multivariable[title/abstract]

chest trauma[title/abstract] AND variate[title/abstract]

chest trauma[title/abstract] AND validat*[title/abstract]

chest trauma[title/abstract] AND predict*[title/abstract]

chest trauma[title/abstract] AND rule*[title/abstract]

chest trauma[title/abstract] AND outcome*[title/abstract]

chest trauma[title/abstract] AND risk*[title/abstract]

chest trauma[title/abstract] AND model*[title/abstract]

chest trauma[title/abstract] AND mortality[title/abstract]

chest trauma[title/abstract] AND survival[title/abstract]

chest trauma[title/abstract] AND complications[title/abstract]

chest injury[title/abstract] AND stratification[title/abstract]

chest injury[title/abstract] AND ROC curve[title/abstract]

chest injury[title/abstract] AND discriminat*[title/abstract]

chest injury[title/abstract] AND c statistic[title/abstract]

chest injury[title/abstract] AND area under the curve[title/abstract]

chest injury[title/abstract] AND AUC[title/abstract]

chest injury[title/abstract] AND calibration[title/abstract]

chest injury[title/abstract] AND indices[title/abstract]

chest injury[title/abstract] AND algorithm[title/abstract]

chest injury[title/abstract] AND multivariable[title/abstract]

chest injury[title/abstract] AND variate[title/abstract]

chest injury[title/abstract] AND validat*[title/abstract]

chest injury[title/abstract] AND predict*[title/abstract]

chest injury[title/abstract] AND rule*[title/abstract]

chest injury[title/abstract] AND outcome*[title/abstract]

chest injury[title/abstract] AND risk*[title/abstract]

chest injury[title/abstract] AND model*[title/abstract]

chest injury[title/abstract] AND mortality[title/abstract]

chest injury[title/abstract] AND survival[title/abstract]

chest injury[title/abstract] AND complications[title/abstract]

thoracic trauma[title/abstract] AND stratification[title/abstract]

thoracic trauma[title/abstract] AND ROC curve[title/abstract]

thoracic trauma[title/abstract] AND discriminat*[title/abstract]

thoracic trauma[title/abstract] AND c statistic[title/abstract]

thoracic trauma[title/abstract] AND area under the curve[title/abstract]

thoracic trauma[title/abstract] AND AUC[title/abstract]

thoracic trauma[title/abstract] AND calibration[title/abstract]

thoracic trauma[title/abstract] AND indices[title/abstract]

thoracic trauma[title/abstract] AND algorithm[title/abstract]

thoracic trauma[title/abstract] AND multivariable[title/abstract]

thoracic trauma[title/abstract] AND variate[title/abstract]

thoracic trauma[title/abstract] AND validat*[title/abstract]

thoracic trauma[title/abstract] AND predict*[title/abstract]

thoracic trauma[title/abstract] AND rule*[title/abstract]

thoracic trauma[title/abstract] AND outcome*[title/abstract]

thoracic trauma[title/abstract] AND risk*[title/abstract]

thoracic trauma[title/abstract] AND model*[title/abstract]

thoracic trauma[title/abstract] AND mortality[title/abstract]

thoracic trauma[title/abstract] AND survival[title/abstract]

thoracic trauma[title/abstract] AND complications[title/abstract]

thoracic injury[title/abstract] AND stratification[title/abstract]

thoracic injury[title/abstract] AND ROC curve[title/abstract]

thoracic injury[title/abstract] AND discriminat*[title/abstract]

thoracic injury[title/abstract] AND c statistic[title/abstract]

thoracic injury[title/abstract] AND area under the curve[title/abstract]

thoracic injury[title/abstract] AND AUC[title/abstract]

thoracic injury[title/abstract] AND calibration[title/abstract]

thoracic injury[title/abstract] AND indices[title/abstract]

thoracic injury[title/abstract] AND algorithm[title/abstract]

thoracic injury[title/abstract] AND multivariable[title/abstract]

thoracic injury[title/abstract] AND variate[title/abstract]

thoracic injury[title/abstract] AND validat*[title/abstract]

thoracic injury[title/abstract] AND predict*[title/abstract]

thoracic injury[title/abstract] AND rule*[title/abstract]

thoracic injury[title/abstract] AND outcome*[title/abstract]

thoracic injury[title/abstract] AND risk*[title/abstract]

thoracic injury[title/abstract] AND model*[title/abstract]

thoracic injury[title/abstract] AND mortality[title/abstract]

thoracic injury[title/abstract] AND survival[title/abstract]

thoracic injury[title/abstract] AND complications[title/abstract]

rib fractures[title/abstract] AND stratification[title/abstract]

rib fractures[title/abstract] AND ROC curve[title/abstract]

rib fractures[title/abstract] AND discriminat*[title/abstract]

rib fractures[title/abstract] AND c statistic[title/abstract]

rib fractures[title/abstract] AND area under the curve[title/abstract]

rib fractures[title/abstract] AND AUC[title/abstract]

rib fractures[title/abstract] AND calibration[title/abstract]

rib fractures[title/abstract] AND indices[title/abstract]

rib fractures[title/abstract] AND algorithm[title/abstract]

rib fractures[title/abstract] AND multivariable[title/abstract]

rib fractures[title/abstract] AND variate[title/abstract]

rib fractures[title/abstract] AND validat*[title/abstract]

rib fractures[title/abstract] AND predict*[title/abstract]

rib fractures[title/abstract] AND rule*[title/abstract]

rib fractures[title/abstract] AND outcome*[title/abstract]

rib fractures[title/abstract] AND risk*[title/abstract]

rib fractures[title/abstract] AND model*[title/abstract]

rib fractures[title/abstract] AND mortality[title/abstract]

rib fractures[title/abstract] AND survival[title/abstract]

rib fractures[title/abstract] AND complications[title/abstract]
