## Supplementary file 2 for "Clinical prediction models for the management of blunt chest trauma in the Emergency Department: a systematic review"

| **Domain** | **Key items** |
| --- | --- |
| **SOURCE OF DATA** | Source of data (e.g., cohort, case-control, randomized trial participants, or registry data) |
| **PARTICIPANTS** | Participant eligibility and recruitment method (e.g., consecutive participants, location, number of centres, setting, inclusion and exclusion criteria) |
|  | Participant description |
|  | Study dates |
| **OUTCOME(S) TO BE PREDICTED** | Definition and method for measurement of outcome |
|  | Was the same outcome definition (and method for measurement) used in all patients? |
|  | Type of outcome (e.g., single or combined endpoints) |
|  | Was the outcome assessed without knowledge of the candidate predictors (i.e., blinded)? |
|  | Were candidate predictors part of the outcome (e.g., in panel or consensus diagnosis)? |
|  | Time of outcome occurrence or summary of duration of follow-up |
| **CANDIDATE PREDICTORS**  **(OR INDEX TESTS)** | Number and type of predictors (e.g., demographics, patient history, physical examination, additional testing, disease characteristics) |
|  | Definition and method for measurement of candidate predictors |
|  | Timing of predictor measurement (e.g., at patient presentation, at diagnosis, at treatment initiation) |
|  | Were predictors assessed blinded for outcome, and for each other (if relevant)? |
|  | Handling of predictors in the modelling (e.g., continuous, linear, non-linear transformations or categorised) |
| **SAMPLE SIZE** | Number of participants and number of outcomes/events |
|  | Number of outcomes/events in relation to the number of candidate predictors (Events Per Variable) |
| **MISSING DATA** | Number of participants with any missing value (include predictors and outcomes) |
|  | Number of participants with missing data for each predictor |
|  | Handling of missing data (e.g., complete-case analysis, imputation, or other methods) |
| **MODEL DEVELOPMENT** | Modelling method (e.g., logistic, survival, neural network, or machine learning techniques) |
|  | Modelling assumptions satisfied |
|  | Method for selection of predictors **for inclusion** in multivariable modelling (e.g., all candidate predictors, pre-selection based on unadjusted association with the outcome) |
|  | Method for selection of predictors **during multivariable modelling** (e.g., full model approach, backward or forward selection) and criteria used (e.g., p-value, Akaike Information Criterion) |
|  | Shrinkage of predictor weights or regression coefficients (e.g., no shrinkage, uniform shrinkage, penalized estimation) |
| **MODEL PERFORMANCE** | Calibration (calibration plot, calibration slope, Hosmer-Lemeshow test) and Discrimination  (C-statistic, D-statistic, log-rank) measures with confidence intervals |
|  | Classification measures (e.g., sensitivity, specificity, predictive values, net reclassification improvement) and whether a-priori cut points were used |
| **MODEL**  **EVALUATION** | Method used for testing model performance: development dataset only (random split of data, resampling methods e.g. bootstrap or cross-validation, none) or separate external validation (e.g. temporal, geographical, different setting, different investigators) |
|  | In case of poor validation, whether model was adjusted or updated (e.g., intercept recalibrated, predictor effects adjusted, or new predictors added) |
| **RESULTS** | Final and other multivariable models (e.g., basic, extended, simplified) presented, including predictor weights or regression coefficients, intercept, baseline survival, model performance measures (with standard errors or confidence intervals) |
|  | Any alternative presentation of the final prediction models, e.g., sum score, nomogram, score chart, predictions for specific risk subgroups with performance |
|  | Comparison of the distribution of predictors (including missing data) for development and validation datasets |
| **INTERPRETATION AND DISCUSSION** | Interpretation of presented models (confirmatory, i.e., model useful for practice versus exploratory, i.e., more research needed) |
|  | Comparison with other studies, discussion of generalizability, strengths and limitations. |
